# Evaluation Oxygen Saturation Monitoring to Detect Pulmonary Edema in Patients with Severe Preeclampsia

**DOI:** 10.1101/2025.05.14.25327559

**Authors:** Adina R. Kern-Goldberger, Bushra Z. Amin, Sindhu K. Srinivas

## Abstract

**Objective:** Pulmonary edema is a feature of preeclampsia with severe features (SPEC), but it is unknown whether oxygen saturation (O2 sat) levels can predict pulmonary edema in patients with SPEC. The purpose of this study is to evaluate and compare O2 sat trends in patients with SPEC with and without pulmonary edema.

**Study Design:** This is a nested case-control study within a retrospective cohort of all patients with SPEC who delivered at a tertiary academic hospital in 2019. Cases were defined as a clinical diagnosis of pulmonary edema (based on imaging findings or clinical concern coupled with empiric treatment with IV diuretic) on postpartum day 0 (PPD0). Controls were patients with SPEC and without pulmonary edema on PPD0. All patients with at least 25 oxygen saturation levels recorded on PPD0 were included. O2 sat trends as well as demographic and clinical features of patients with and without pulmonary edema were compared.

**Results:** 238 patients in total were included. Five patients (2.1%) were diagnosed with pulmonary edema on PPD0. There were no significant differences in demographic or major obstetric characteristics of patients with and without pulmonary edema. There were also no significant differences between groups in the number of patients with at least one abnormal O2 sat, with a large volume of abnormal O2 sat values in both groups. Significant differences were seen in the minimum oxygen saturation level recorded (92 v. 89, p < 0.04) and the overall percent of abnormal values (7.0% abnormal values without pulmonary edema compared to 46.0% with pulmonary edema, p < 0.01).

**Conclusions:** Individual O2 sat values are poorly predictive of pulmonary edema in patients at risk, but trends may be more prognostic. Algorithms that employ real-time data trending may position O2 sat as a better surveillance tool for pulmonary edema in patients with SPEC.

## Objective

Pulmonary edema is a complicating feature of preeclampsia with severe features (SPEC), but little is known about whether oxygen (O2) saturation levels can actually predict pulmonary edema in patients at risk.^1^ Oxygen desaturation is a clinical sign of pulmonary edema, but intrapartum O2 monitoring can be subject to measurement error, and downtrends in O2 saturation may not antecede clinically significant pulmonary edema with enough forewarning to facilitate a preventative or early response. Delays in diagnosis have been identified as a major potential contributor to preventable severe maternal morbidity (SMM) associated with preeclampsia.^2^ The purpose of this study, therefore, is to evaluate and compare O2 saturation trends in patients with SPEC with and without pulmonary edema.

## Study Design

This is a nested case-control study within a retrospective cohort of all patients with SPEC diagnosed on or before PPD0 who delivered at a tertiary academic hospital in 2019. Cases were defined by clinical diagnosis of pulmonary edema (based on imaging findings or documented clinical concern coupled with empiric treatment with intravenous diuretic) on postpartum day 0 (PPD0). Controls were all patients with severe preeclampsia and without pulmonary edema on PPD0. All patients with at least 25 individual oxygen saturation levels recorded on PPD0 were included. Patients with other potential etiologies of low O2 saturation (acute pulmonary embolism, pneumonia, cardiomyopathy) were excluded. O2 saturation values and patterns as well as demographic and clinical features of patients with and without pulmonary edema were compared with chi-square, Student’s t-test, and Wilcoxon rank-sum tests as appropriate. Abnormal O2 saturation was defined as < 95%. This study was considered exempt by the Institutional Review Board (Protocol # 848479).

## Results

In total, 238 patients with severe preeclampsia were included. Five patients (2.1%) were diagnosed with pulmonary edema on PPD0. Patients with pulmonary edema had lower delivery BMI and were more likely to have had an earlier diagnosis of SPEC [median 26 vs. 35 weeks] as well as an unplanned or emergency cesarean (Supplemental Table 1). They were also more likely to be admitted to the intensive care unit (ICU) or have SMM without transfusion (Supplemental Table 2). There were no differences between groups in the number of patients with at least one abnormal O2 saturation value, and PPD0 O2 trends for all patients are depicted in the Figure 1, demonstrating a large volume of abnormal values for both groups. However, mean (95.4% [SD 1.45] for pulmonary edema vs. 97.9% [SD 0.81] without [p < 0.01]), median (95% [IQR 94-96] with pulmonary edema vs. 99% [IQR 98-100] without [p < 0.01]), and absolute minimum O2 saturation level recorded were all significantly lower for patients with pulmonary edema, while median percent of abnormal values recorded was higher (46% for pulmonary edema [IQR 44-54] vs. 6.98% [IQR 2-18] without [p < 0.01]) (Table 1).

**Table 1.** Oxygen Saturation Parameters in Patients with and without Pulmonary Edema.

|  | **No Pulmonary**<br>**Edema**<br>**N = 233** | **Pulmonary Edema**<br>**N = 5** | **P Value** |
| --- | --- | --- | --- |
| **Mean (SD)** | 97.9 (1.45) | 95.4 (0.81) | **< 0.01** |
| **Median (IQR)** | 99 (98, 100) | 95 (94, 96) | **< 0.01** |
| **Minimum (median, IQR)** | 92 (90-94) | 89 (88-89) | **0.04** |
| **Any abnormal value < 95% (N, %)** | 183 (78.54) | 5 (100) | 0.25 |
| **Percent of abnormal values < 95%**<br>**(median, IQR)** | 6.98 (2, 18) | 46 (44, 54) | **< 0.01** |

**Figure 1.**
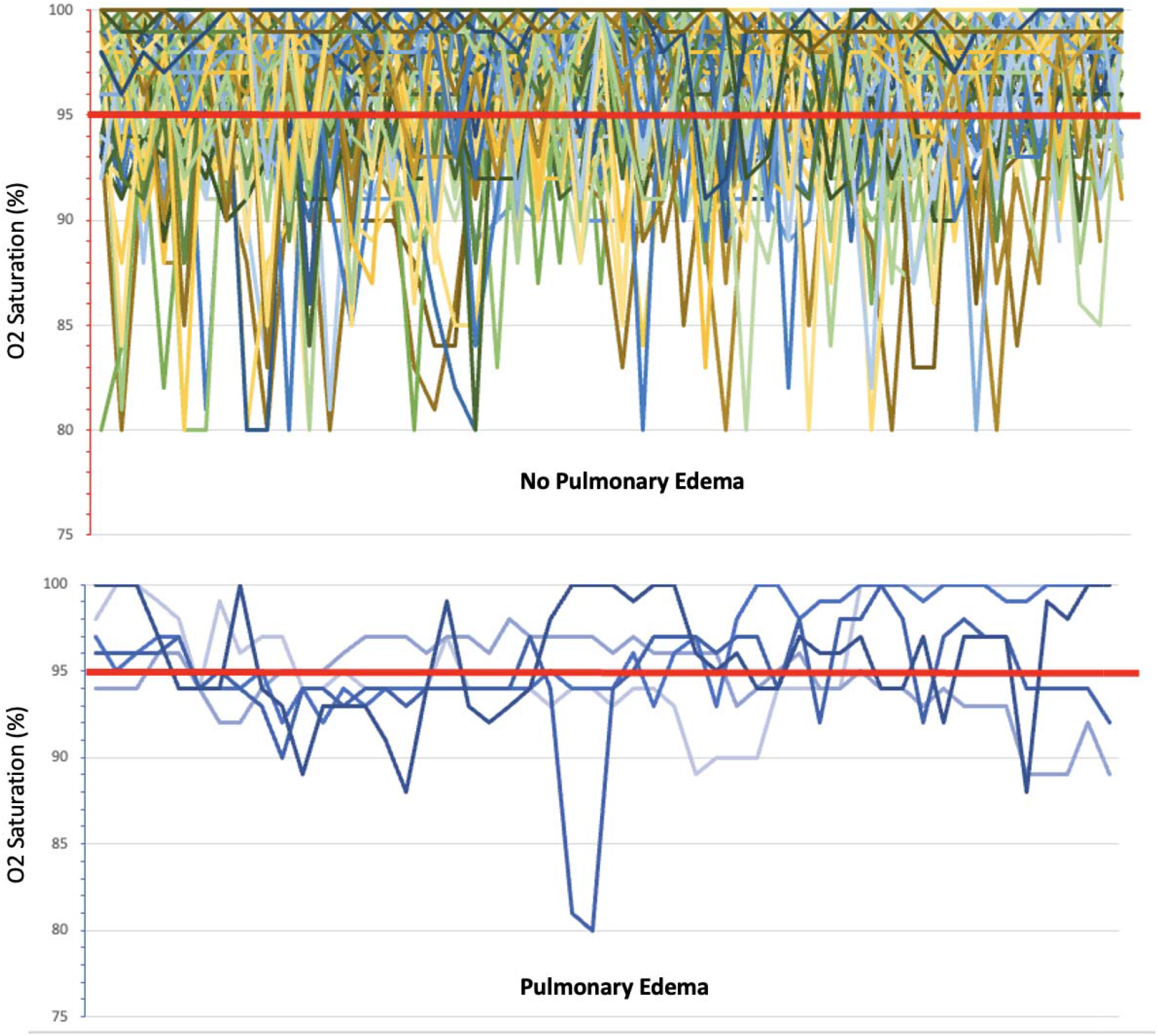
Individual Oxygen Saturation Trends of Patients with and without Pulmonary Edema

## Conclusion

These data suggest that overall, individual O2 sat values are poorly predictive of pulmonary edema in patients at risk, which may generate signal saturation and alarm fatigue. ^3^ A median of nearly 7% of all O2 saturations measured for patients without pulmonary edema were abnormal. While O2 saturation values were generally more abnormal in patients with pulmonary edema, the specific diagnostic yield of traditional O2 saturation monitoring is difficult to determine. O2 saturation trends may be more predictive of pulmonary edema than single measurements, and algorithms that employ real-time data trending in the electronic medical record and machine learning may position O2 saturation as a better surveillance tool for pulmonary edema in patients with SPEC, as has been demonstrated for other obstetric morbidities such as hemorrhage.^4^

Though the sample of pulmonary edema patients in this study is small, these data provide granular, patient level vital sign data from a cohort of patients at highest risk for intrapartum or immediate postpartum preeclampsia-related pulmonary edema, which, though uncommon, may be an important source of preventable morbidity. Enhancing early detection of pulmonary edema has the potential to reduce related SMM, and further research should explore the clinical effectiveness and implementation of O2 saturation trending and machine learning to further this goal. There may also be value in introducing targeted surveillance for patient at highest risk of pulmonary edema, such as those with preterm SPEC and those undergoing urgent or emergent cesarean delivery, as identified in this study.

## Data Availability

All data produced in the present study are available upon reasonable request to the authors.

---

**Supplemental Table 1.**
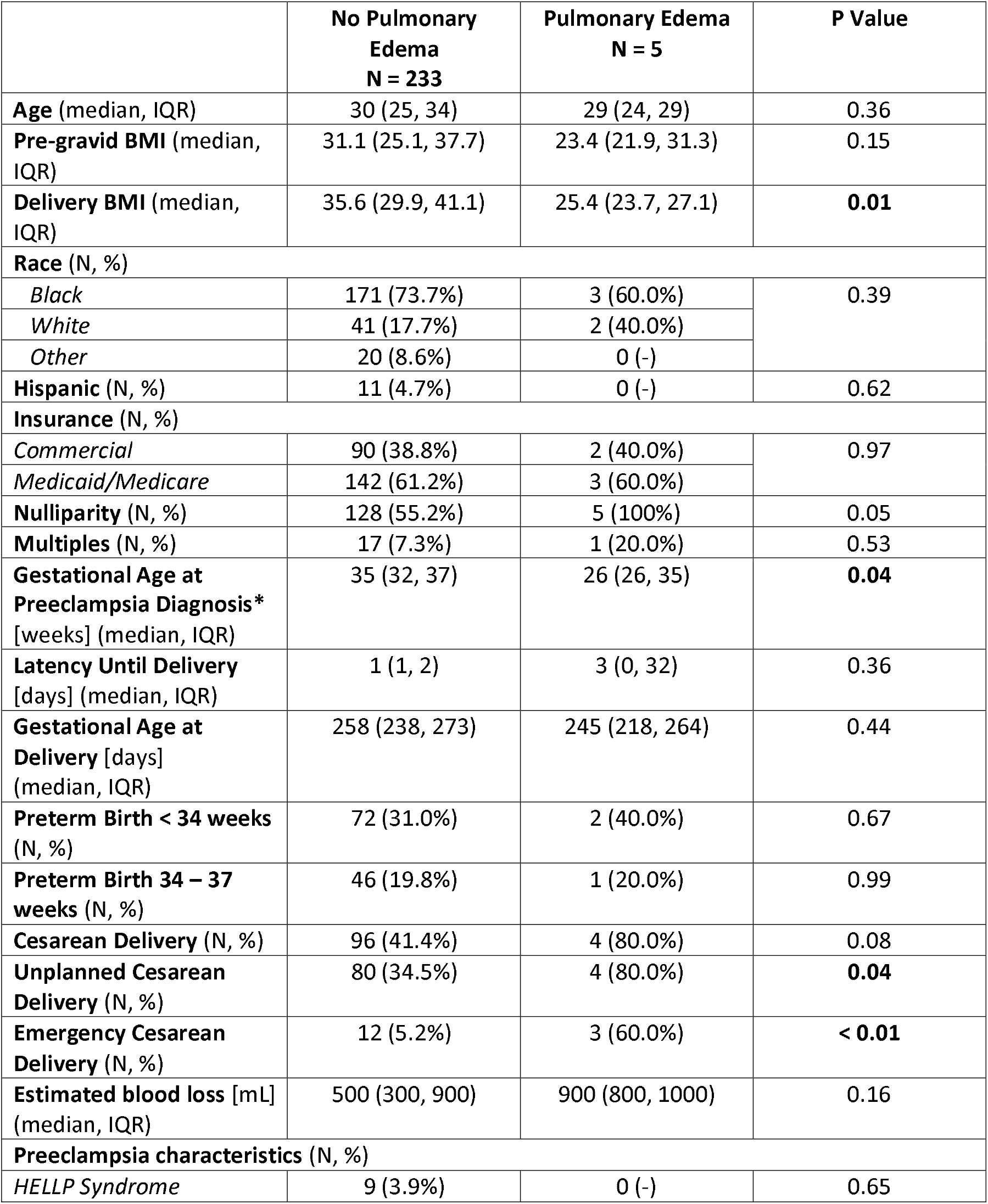
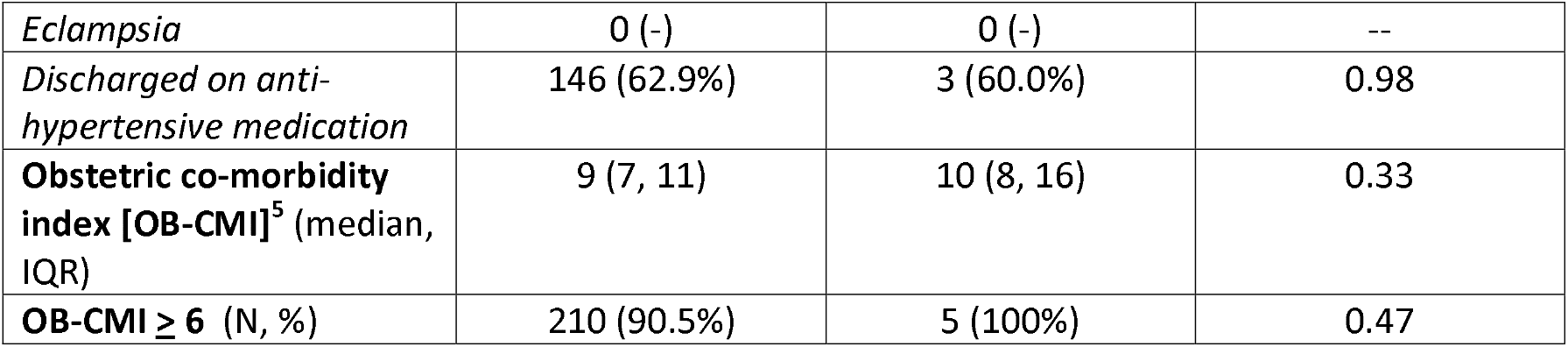
Demographics and Obstetric Characteristics in Patients with and without Pulmonary Edema.

**Supplemental Table 2.** Maternal Outcomes in Patients with and without Pulmonary Edema.

|  | **No Pulmonary Edema**<br>**N = 233** | **Pulmonary Edema**<br>**N = 5** | **P Value** |
| --- | --- | --- | --- |
| **Total length of stay**<br>(median, IQR) | 4 (3, 5) | 7 (4, 9) | 0.07 |
| **Postpartum length of stay** (median, IQR) | 3 (2, 4) | 4 (4, 4) | 0.05 |
| **Postpartum length of stay ≥ 5 days (N, %)** | 13 (5.6%) | 1 (20.0%) | 0.18 |
| **ICU admission (N, %)** | 4 (1.7%) | 2 (40.0%) | **< 0.01** |
| **Readmission (N, %)** | 7 (3.0%) | 0 (0%) | 0.69 |
| **SMM\* with Transfusion (N, %)** | 46 (19.8%) | 2 (40.0%) | 0.27 |
| **SMM \*without Transfusion (N, %)** | 16 (6.9%) | 2 (40.0%) | **< 0.01** |

\*Severe maternal morbidity as defined by the Centers for Disease Control ([https://www.cdc.gov/reproductivehealth/maternalinfanthealth/smm/severe-morbidity-](https://www.cdc.gov/reproductivehealth/maternalinfanthealth/smm/severe-morbidity-ICD.htm) [ICD.htm](https://www.cdc.gov/reproductivehealth/maternalinfanthealth/smm/severe-morbidity-ICD.htm))

